# Diagnostic accuracy of Chest X-Ray Computer Aided Detection software and blood biomarkers for detection of prevalent and incident tuberculosis in household contacts followed up for 5 years

**DOI:** 10.1101/2024.06.30.24309731

**Authors:** Liana Macpherson, Sandra V. Kik, Matteo Quartagno, Francisco Lakay, Marche Jaftha, Nombuso Yende, Shireen Galant, Saalikha Aziz, Remy Daroowala, Richard Court, Arshad Taliep, Keboile Serole, Rene T. Goliath, Nashreen Omar Davies, Amanda Jackson, Emily Douglass, Bianca Sossen, Sandra Mukasa, Friedrich Thienemann, Taeksun Song, Morten Ruhwald, Robert J. Wilkinson, Anna K. Coussens, Hanif Esmail Imaging of TB household contacts group

## Abstract

**Background:** WHO Tuberculosis (TB) screening guidelines recommend computer-aided detection (CAD) software for chest radiograph (CXR) interpretation. However, studies evaluating their diagnostic and prognostic accuracy are limited.

**Methods:** We conducted a prospective cohort study of household TB contacts in South Africa. Participants all underwent baseline CXR and sputum investigation (routine [single spontaneous] and enhanced [additionally 2-3 induced] sputum investigation and passive and active follow-up for incident TB. CXR were processed comparing 3 CAD softwares (CAD4TBv7.0, qXRv3.0.0, and Lunit INSIGHT CXR 3.1.4.111). We evaluated their performance to detect routine and enhanced prevalent, and incident TB, comparing the performance to blood-based biomarkers (Xpert MTB host-response, Erythrocyte Sedimentation Rate, C-Reactive Protein, QuantiFERON) in a subgroup.

**Findings:** 483 participants were followed-up for 4.6 years (median). There were 23 prevalent (7 routinely diagnosed) and 38 incident TB cases. The AUC ROC to identify prevalent TB for CAD4TB, qXR and Lunit INSIGHT CXR were 0.87 (95% CI 0.77-0.96), 0.88 (95% CI 0.79-0.97) and 0.91 (95% CI 0.83-0.99) respectively. >30% with scores above recommended CAD thresholds who were bacteriologically negative on routine baseline sputum were subsequently diagnosed by enhanced baseline sputum investigation or during follow-up. The AUC performance of baseline CAD to identify incident cases ranged between 0.60-0.65. The diagnostic performance of CAD for prevalent TB was superior to blood-based biomarkers.

**Interpretation:** Our findings suggest that the potential of CAD-CXR screening for TB is not maximised as a high proportion of those above current thresholds but with a negative routine confirmatory sputum have true TB disease that may benefit intervention.

**Funding:** UKRI-MRC

**Summary:** We found that the diagnostic accuracy of CAD-CXR to identify prevalent TB cases in household TB contacts was high but >30% with scores above recommended CAD thresholds who were bacteriologically negative on routine testing baseline were subsequently diagnosed suggest that the potential of CAD-CXR screening is not maximised.

## Introduction

The World Health Organisation (WHO) estimated that there are up to 4 million people living with undiagnosed tuberculosis (TB) (1). Active case finding aims to proactively identify individuals with pulmonary TB not seeking healthcare to enable earlier treatment, thus reducing morbidity, mortality, and transmission. As 50% of undiagnosed bacteriologically positive cases in the community are symptom screen negative, and it is increasingly evident that CXR is more effective to identify the undiagnosed TB cases (2–7). The sensitivity of CXR-based screening for active TB is emphasised in the updated 2021 WHO screening guidelines, which also endorse for the first time the use of computer aided detection (CAD) software for automated radiographic interpretation in those aged >15 years (8). The Stop TB Partnership’s *Global Plan to Stop TB* also highlights the critical role active case finding in reducing TB incidence (9).

CAD software use trained deep learning algorithms and artificial intelligence to interpret CXR for signs of TB with several studies having reported equivalent accuracy compared to human readers (8,10–12). However, high quality, prospective studies evaluating the diagnostic accuracy of such software are limited, particularly in the screening setting and with comprehensive sampling and follow-up for TB (13,14).

Confirmatory bacteriological testing is often done by assessment of a single spontaneously produced sputum sample with molecular detection of *Mycobacterium tuberculosis* (*Mtb*) DNA (e.g. Xpert MTB/RIF) (8). This approach is insensitive as not all those with a positive triage test are able to expectorate sputum for confirmatory testing. In a recent research study evaluating CAD, only 29% were able to produce a valid sputum (15). Bacteriologically confirmation increases with multiple samples, sputum induction and/or use of culture, but such enhanced measures are not typically performed during routine screening.

Those with CXR abnormalities without sputum confirmation are at high risk of disease progression, a recent meta-analysis demonstrated that individuals with CXR changes suggestive of TB but with negative sputum bacteriology subsequently have a 10% risk per year to being diagnosed with bacteriologically positive TB disease (16). However, the majority of the contributory studies were historical, pre-HIV epidemic, and used conventional CXR and no contemporary studies have evaluated this risk utilising digital CXR and CAD approaches.

In addition, blood tests that are predictive of future TB risk have recently been proposed as a priority for development with blood based transcriptional markers now in late stages of development. Such blood-based assays could potentially be used alongside CXR screening for prevalent TB disease to identify individuals with future TB risk. However, evaluation of these tests in a screening population alongside or in combination with CXR has not previously been undertaken.

In this study we screened household contacts (HHC) of rifampicin-resistant (RR) TB patients by digital CXR and undertook long term follow-up for development of TB disease in the absence of preventive therapy. Our aims were to: (1) Evaluate the diagnostic accuracy of three CAD software packages against microbiological reference standards to detect prevalent and incident pulmonary TB on baseline CXR. (2) Determine the sensitivity and specificity of CAD software using the recommended threshold scores. (3) Compare and combine CAD scores with a blood tests (Xpert MTB host-response, Erythrocyte Sedimentation Rate (ESR), C-Reactive Protein (CRP), QuantiFERON) to detect prevalent and incident TB.

## Methods

### Setting and participants

This prospective cohort study was conducted in Khayelitsha, South Africa. Recruitment of household contacts (HHC) of at least rifampicin resistant (RR) TB index cases took place between November 2014 and September 2017 with follow up until May 2021 (17) (see Supplementary method for details). Ethical approval for this study was received from: University of Cape Town (449/2014), Boston University (H-35831), Rutgers University (Pro2018001966), NIH (DMID 16-0112) and University College London (19219/001). This report follows the STARD guidelines for diagnostic accuracy studies (18).

### Procedures

Eligible individuals were HHC aged ≥18 years. At screening participants underwent medical history, physical examination, HIV, TB symptoms screen (unexplained cough for ≥ 2 weeks, fever, night sweats and weight loss), and a digital CXR. All participants were extensively investigated for TB at baseline regardless of symptoms, with 1 attempted spontaneous, spot, sputum sample followed by 2-3 induced sputum samples to a total of 3 samples. Sputum induction was by nebulisation of 3% hypertonic saline. Samples were processed in the accredited laboratories where auramine sputum smear, Xpert MTB/RIF and mycobacterial growth indicator tube (MGIT) liquid TB culture were performed according to local standard operating procedures. Female participants of child-bearing potential underwent urinary pregnancy testing and if positive were excluded from the study.

As part of a nested study, participants, who were HIV-uninfected and asymptomatic, additionally consented to also undergo blood sampling for a biomarker sub-study (17). Full details of inclusion and exclusion criteria can be found in the Supplementary Methods. Blood samples were taken for analysis, including full blood count (FBC), serum C-reactive protein (CRP), erythrocyte sedimentation rate (ESR), QuantiFERON Gold (QFT-Gold) (Qiagen) and Tempus Tubes (Applied Biosystems) stored at −80°C to preserve blood RNA for transcriptomic analysis including the 3-gene RNA Xpert MTB Host Response [MTB-HR] (Cepheid, Sunnyvale, Ca, USA). Further details are in the Supplementary Methods.

No participants received preventive therapy as they were contacts of RR-TB in line with national and some international guidelines at the time of the study recommending close follow-up. (19) Participants who were bacteriologically positive or with clinical concern of TB were referred to the statuory TB clinic where the decision to start TB treatment was determined.

### Follow up

Participants were asked to attend the clinic for assessment if they developed TB symptoms. All participants were then invited for systematic re-screening between 24 and 36 months irrespective of symptoms with 3 sputum samples taken (induced if needed) sent for smear, Xpert MTB/RIF and culture. To ensure that all episodes of treated TB within the Western Cape Province were captured, participants consented to access to their health service records via the Provincial Health Data Centre (PHDC) and clinical medical records (20). This search took place on 19^th^ May 2021.

### Chest radiograph reading

Posterior-anterior CXR in full inspiration were performed using a digital X-Ray machine (Phillips Essenta DR). All CXR were reported by a medical officer blinded to clinical details and microbiological outcomes and were classified as abnormal – TB related, abnormal – not TB related and normal.

Three commercially available CAD software: CAD4TB version 7.0 (CAD4TBv7, Delft Imaging, ‘s-Hertogenbosch Netherlands), qXR version 3.0.0 (qXRv3, qure.ai, Mumbai, India) and Lunit INSIGHT CXR version 3.1.4.111 (Lunit INSIGHT CXRv3, Lunit, Seoul, South Korea) (see Supplementary Methods for details and manufacturer recommended threshold scores).

### Reference Standards

For the determination of the diagnostic accuracy a case of prevalent TB was defined as at least 1 baseline sputum sample culture and/or Xpert MTB/RIF positive for *Mtb* where the participant was treated for TB. We created subgroups of participants with prevalent disease to reflect cases that would be identified through routine screening (routine prevalent) and those who would only be identified through more intensive investigation (enhanced prevalent). Routine prevalent cases were defined as those cases initiated on TB treatment in whom *Mtb* was detected using Xpert MTB/RIF on the first single spontaneously produced baseline sputum sample, these are cases that would be detected by current screening practices in South Africa (21). Enhanced prevalent cases were defined as those initiated on TB treatment in whom *Mtb* was detected in any other baseline sputum sample by either Xpert MTB/RIF or culture. Incident TB was defined as cases initiated on TB treatment in whom at least 1 follow up sputum sample culture and/or Xpert MTB/RIF was *Mtb* positive or where a clinician independent to the study made a clinical decision to start TB treatment. The inclusion of clinically diagnosed TB was done to capture participants diagnosed and treated elsewhere during the follow up period ascertained through the PHDC records. We classified participants as not having TB if all baseline and follow up samples were negative for *Mtb*, and they were not initiated on TB treatment.

### Statistical analysis

Sample size was determined by the parent study (17). Area under the receiver operator curve (AUC ROC) was calculated to evaluate diagnostic performance for the CAD software and other biomakers. We calculated the sensitivity and specificity of CAD software using the manufacturers pre-specified or commonly used thresholds. Analyses were also performed for a number of pre-specified subgroups chosen because of known associations with TB risk, clinical presentation, and plausibility that they might affect CXR findings: people living with HIV (PLHIV), participants with a history of previous TB, and smokers. For missing data, participants were excluded from analysis if any one of the CAD software did not provide a score due to error, or data was incomplete for the baseline sputum results. Statistical analyses were done with R version 4.0.5 (2021-03-31).

## Results

983 HHC of RR-TB were identified, of whom 511 eligible adult participants consented and were screened for TB. 483 participants were included in the analysis following exclusions (Figure 1). Median age was 33 years, 308 (61%) were female, 109 (23%) had a history of previous TB, and 136 (28%) were PLHIV (Table 1). For those PLHIV 62.5% were on antiretroviral therapy (ART) and a recent CD4 count was available for 54 participants (median 413.5/mm^3^ (IQR 236-562)). The participants were followed up for a median of 4.6 years (IQR 3.9-5.2 years) 247 HIV-uninfected, participants without TB symptoms met the inclusion criteria to undergo biomarker blood sampling.

**Figure 1:**
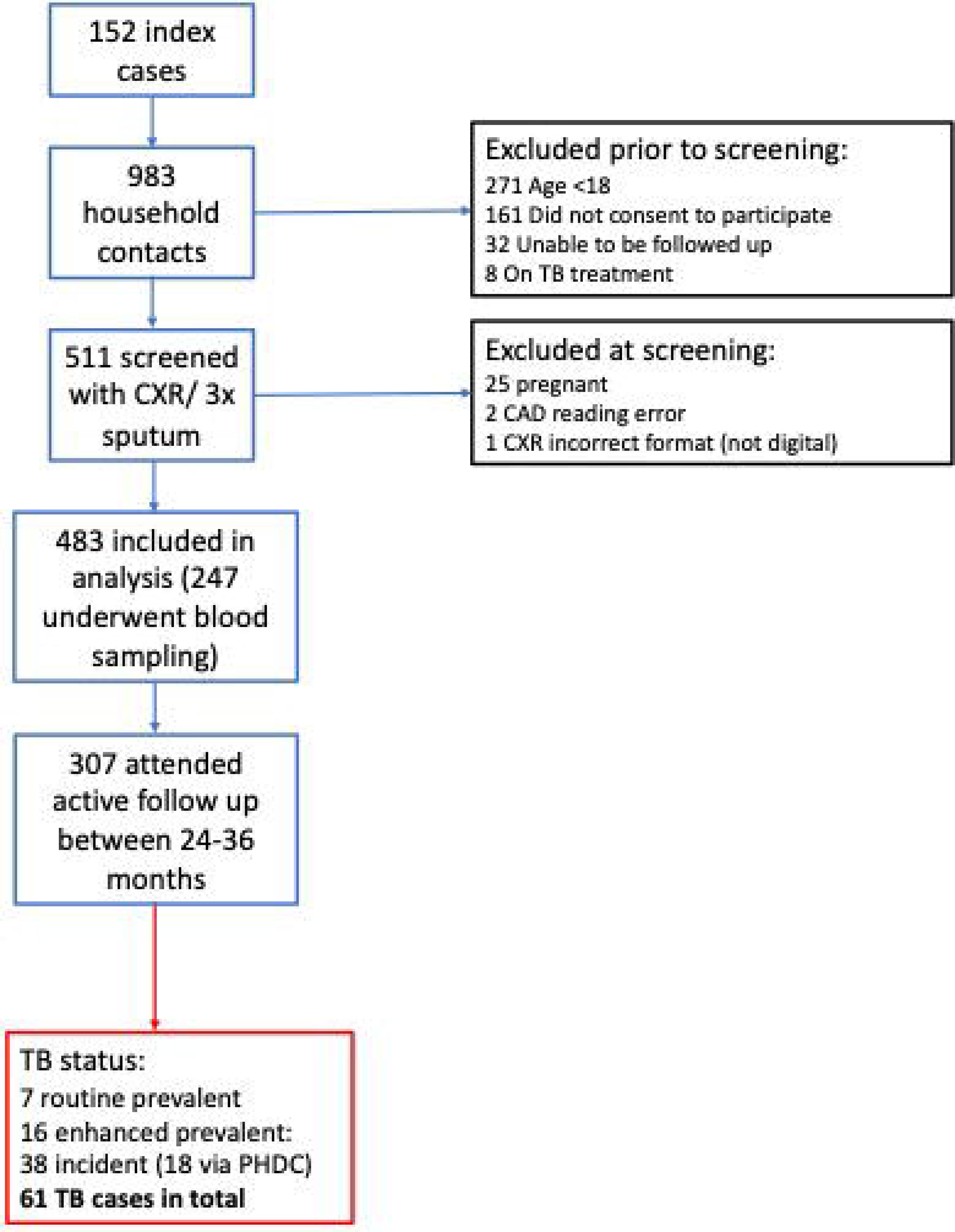
Study population. Showing the details of participants who were included in this analysis including a description of TB disease status.

**Table 1:** Baseline characteristics. Detailing the participants demographic data, sputum samples, and scores from the computer aided detection (CAD) software for baseline chest radiograph interpretation. This information is displayed for all participants, those with routinely diagnosed prevalent Tuberculosis (TB), enhanced prevalent TB, and incident TB.

### Yields of different screening strategies

Of the 483 participants, at baseline screening, 60% produced a spontaneous sputum sample, 96% an induced sample, and 84% had at least 3 sample results available (Table 1). 23 (4.7%) had bacteriologically confirmed TB infection at baseline (prevalent TB), 7 (30%) of these were routine prevalent cases, and the remaining 16 were enhanced prevalent cases.

A further 38 (7.9%) cases of incident TB, including 18 diagnosed elsewhere and captured via PHDC, were identified during follow-up. Thirteen (34%) were diagnosed within 12 months, and 18 (47%)) between 24-36 months, median time to diagnosis was 24 months (IQR 10-34). Six incident cases were clinically diagnosed, but not bacteriologically confirmed.

Fifty-one (11%) participants reported at least 1 TB symptom at baseline, of these, 8 were confirmed TB at baseline (35% of the 23 prevalent cases). Of the 23 prevalent cases, 8 (35%) were smear positive, 14 (61%) were Xpert MTB/RIF positive and 20 (87%) were culture positive in baseline samples (Supplementary Tables 1a and b). Of the 38 incident cases 79% reported TB symptoms at the time of diagnosis and 3 had extra-pulmonary disease.

Participants in this study underwent more intensive screening investigations compared to international screening recommendations (22). We explored the yields of hypothetical screening strategies: a) Positive symptom screen followed by Xpert MTB/RIF on a spontaneously produced sputum sample and b) positive CXR (human read) followed by Xpert MTB/RIF on a spontaneously produced sputum sample. These strategies detected 4/23 (17%) and 5/23 (22%) of the prevalent TB cases in this cohort respectively. (Figure 2).

**Figure 2:**
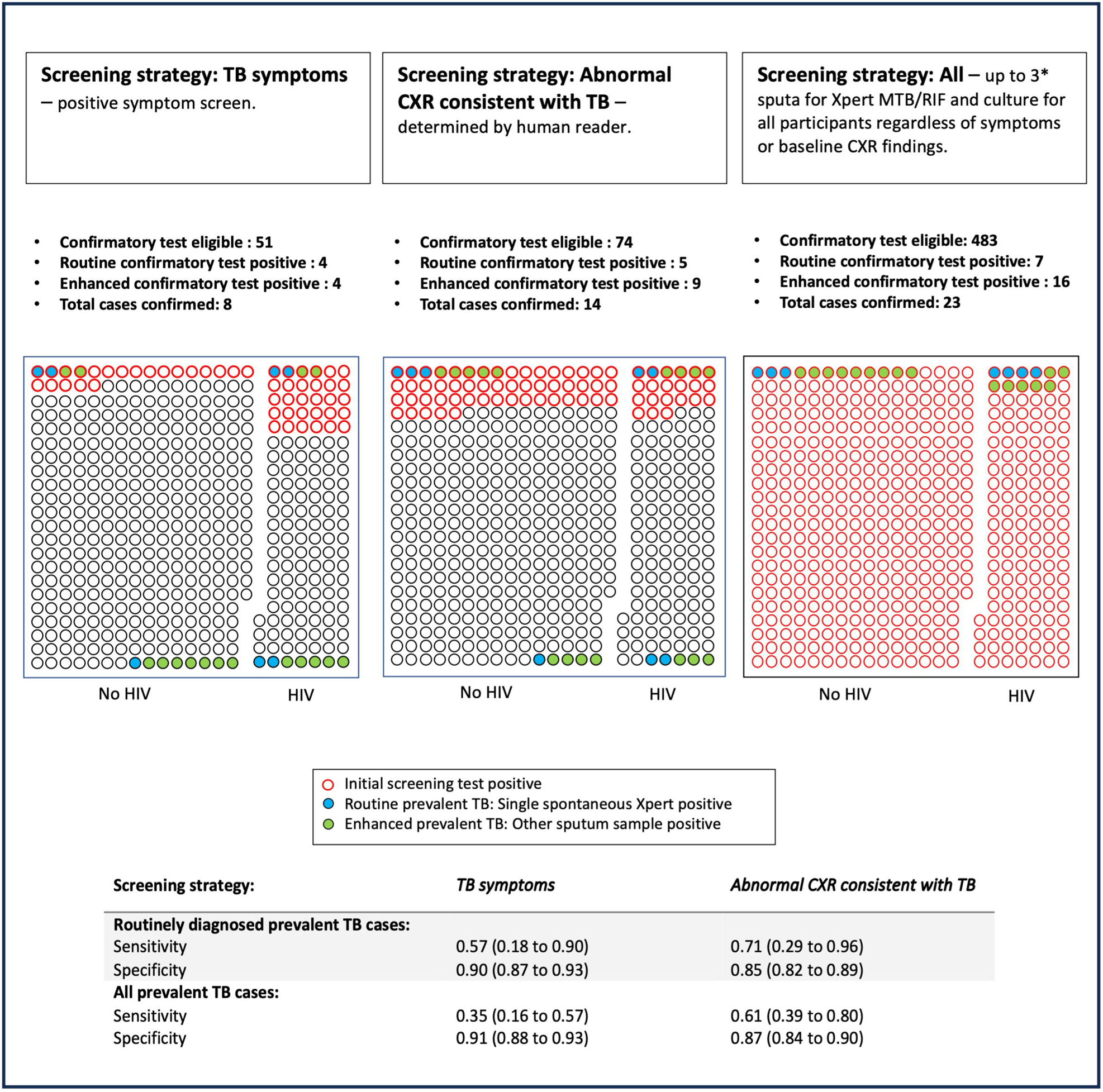
Yields of three different TB screening strategies. In this representation of the yields of these screening each dot represents one study participant. The colour coding of the participants represents Tuberculosis (TB) disease status as defined by bacteriological investigation. Participants have also been grouped by HIV status to show the relative representation of TB in each population. The figure is accompanied by a table which shows the sensitivity and specificity of the different screening approaches (the numbers represent proportions and 95% confidence intervals.

### CAD results

All CAD software performed with highest accuracy for identifying prevalent TB and there were no statistically significant differences between CAD systems for any of the comparisons. The AUC ROC were 0.85-0.95 for identifying routine prevalent and 0.85-0.89 for identifying enhanced prevalent cases (Figure 3). The CAD software performed less well at identifying participants with incident TB on baseline CXR with AUC ROC of 0.60-0.65. For incident disease, there was no substantial difference in the accuracy to detect TB occurring between 1-12 months, 13-24 months or over 24 months after study entry (Supplementary Figure 1).

**Figure 3:**
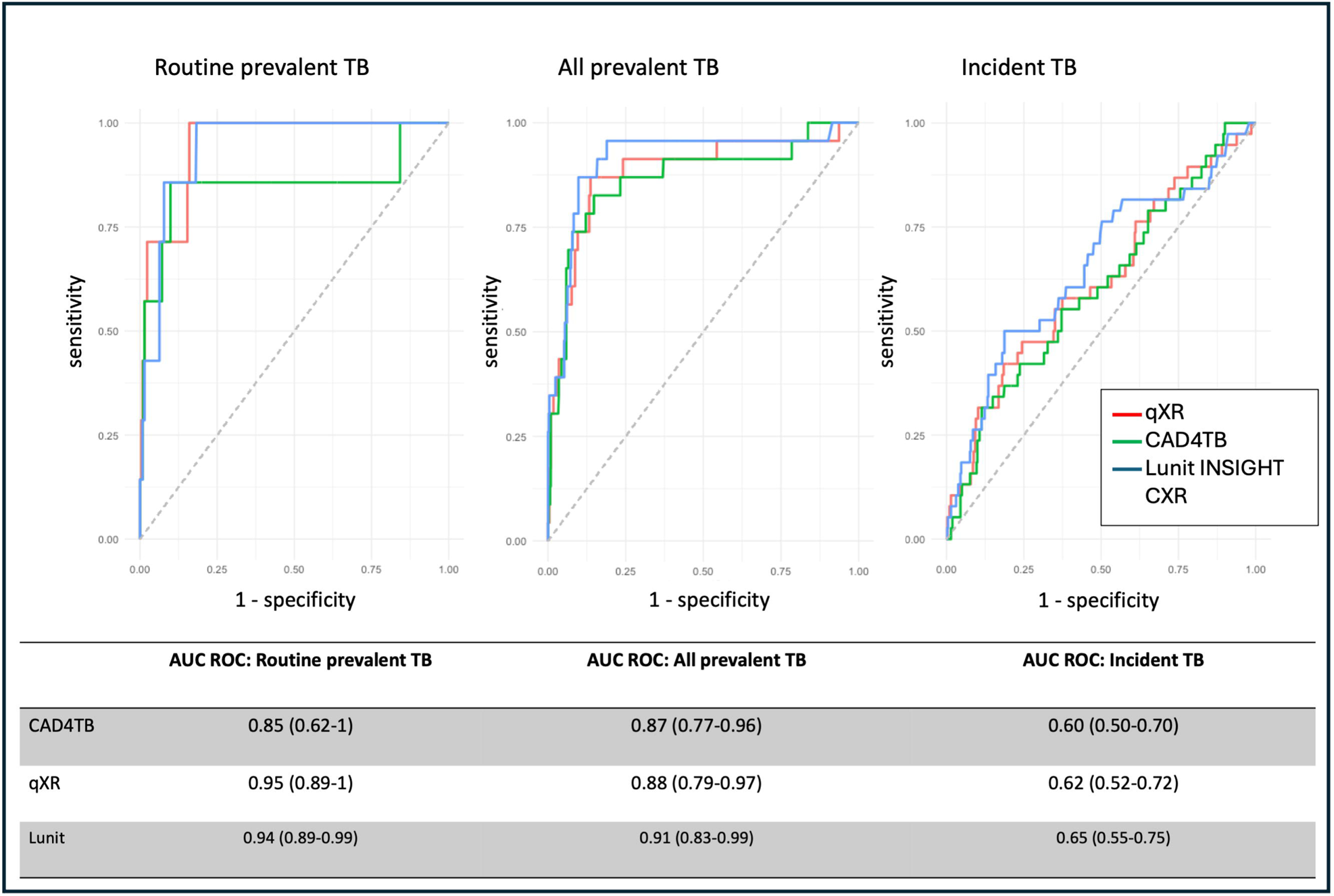
Performance of three different computer-aided detection (CAD) software for the detection of prevalent and incident Tuberculosis (TB) cases. The CAD software evaluated were CAD4TB version 7.0 (CAD4TBv7, Delft Imaging, ‘s-Hertogenbosch Netherlands), qXR version 3.0.0 (qXRv3, qure.ai, Mumbai, India) and Lunit INSIGHT CXR version 3.1.4.111 (Lunit INSIGHT CXRv3, Lunit, Seoul, South Korea). Routine prevalent TB was defined as those initiated on TB treatment in whom Mycobacterium tuberculosis (Mtb) was detected using Xpert MTB/RIF on the first single spontaneously produced baseline sputum sample, all prevalent TB was defined as those initiated on TB treatment in whom at Mtb was detected using Xpert MTB/RIF and/or culture on any other baseline sputum sample, and incident TB was defined as those cases initiated on TB treatment in whom Mtb was detected in at least one follow up sputum sample by Xpert MTB/RIF and/or culture or where the initiation of TB treatment was based on clinical grounds. Accuracy was measured using area under receiver operator curve (AUC ROC). The numbers represent proportion (95% confidence interval).

All CAD systems detected prevalent TB with significantly greater accuracy in participants with no history of previous TB. There was also a trend to better performance in those who were not living with HIV (see Supplementary Table 2).

Using the manufacturer recommended thresholds, the sensitivity/specificity to detect routine prevalent TB cases was 0.71/0.91 for CAD4TB, 0.72/0.93 for qXR and 0.86/0.83 for Lunit INSIGHT CXR. The sensitivity/specificity to detect all prevalent cases was 0.7/0.93 for CAD4TB, 0.57/0.94 for qXR and 0.87/0.86 for Lunit INSIGHT CXR (see Supplementary Table 3).

Between 8% and 18% of participants had CAD scores above recommended threshold and of those 7-13% were diagnosed as prevalent cases with routine sampling. However, notably 30-35% of those above threshold, not diagnosed routinely, were subsequently diagnosed and treated for TB either as prevalent cases identified through enhanced sampling or as incident cases (i.e. cannot be considered as a “false positive”). For participants above threshold not diagnosed or treated for TB over 5 years all had a previous history of TB with CAD4TB and qXR, and 87% (46/53) with Lunit INSIGHT CXR (Figures 4 and 5).

**Figure 4:**
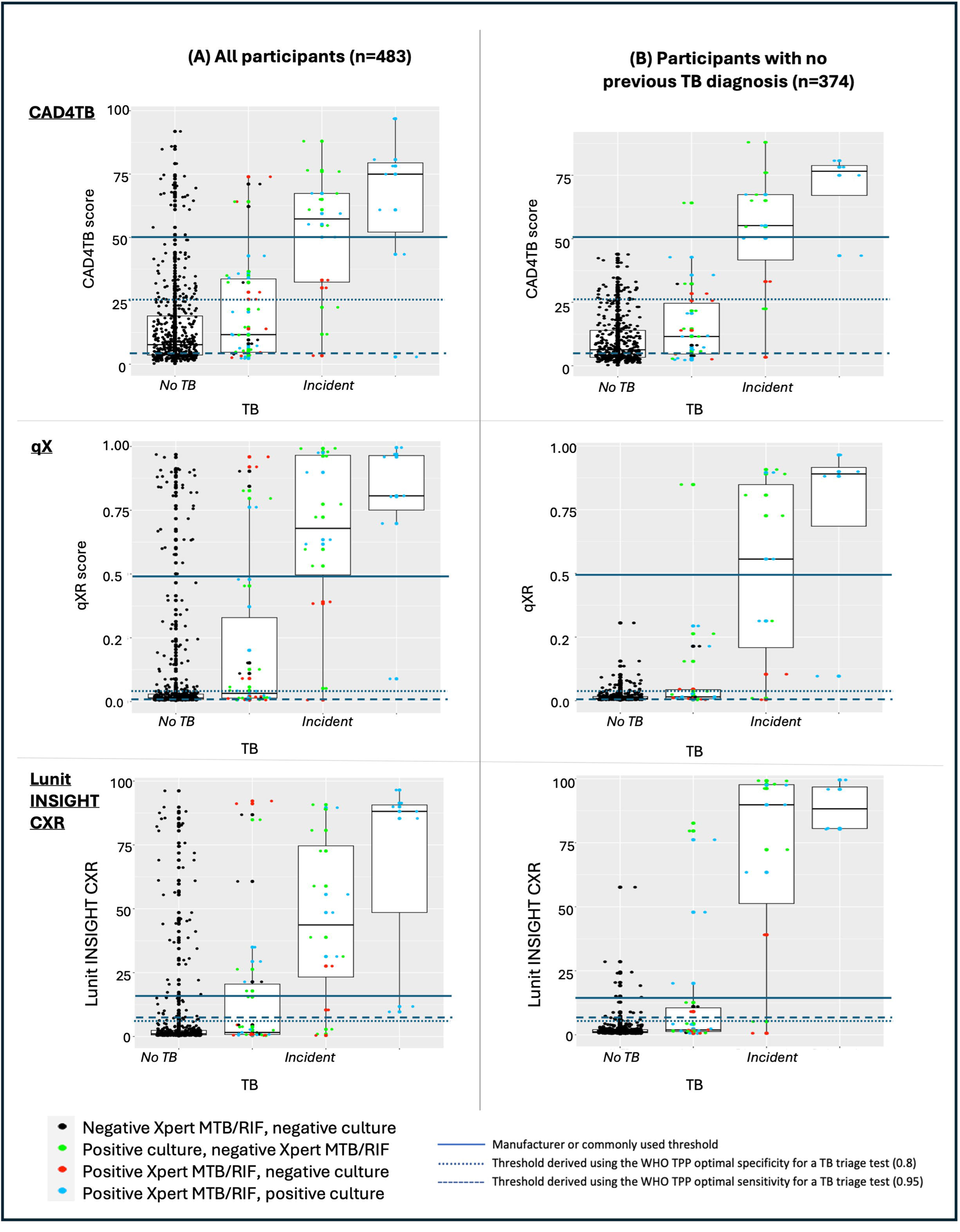
Representation of study participants above and below the manufacturer or commonly used threshold for each CAD software (CAD4TB, qXR and Lunit INSIGHT CXR) by TB status. This figure provides a visual representation of the distribution of computer-aided detection software scores for each software used (CAD4TB v7.0, qXR v3.0.0, Lunit INSIGHT CXR v3.1.4.111), broken down by TB disease status. Routine prevalent TB was defined as those initiated on TB treatment in whom Mycobacterium tuberculosis (Mtb) was detected using Xpert MTB/RIF on the first single spontaneously produced baseline sputum sample, all prevalent TB was defined as those initiated on TB treatment in whom at Mtb was detected using Xpert MTB/RIF and/or culture on any other baseline sputum sample, and incident TB was defined as those cases initiated on TB treatment in whom Mtb was detected in at least one follow up sputum sample by Xpert MTB/RIF and/or culture or where the initiation of TB treatment was based on clinical grounds. Each dot represents a study participant, and the colour coding of the dot represents the bacteriological status of that participant. For example, some incident cases are represented by black dots indicating that these participants were bacteriologically negative, these represent the cases of incident TB that were diagnosed clinically. Horizontal lines have been added to represent three thresholds – the threshold above which the score is consistent with TB infection as recommended by the manufacturer or commonly used in practice, the threshold derived by using the WHO target product profile (TPP) optimal specificity for a TB triage test, and the threshold derived by using the WHO target product profile (TPP) optimal sensitivity for a TB triage test. The latter two thresholds were generated using all study participants/all prevalent cases of TB.

**Figure 5:**
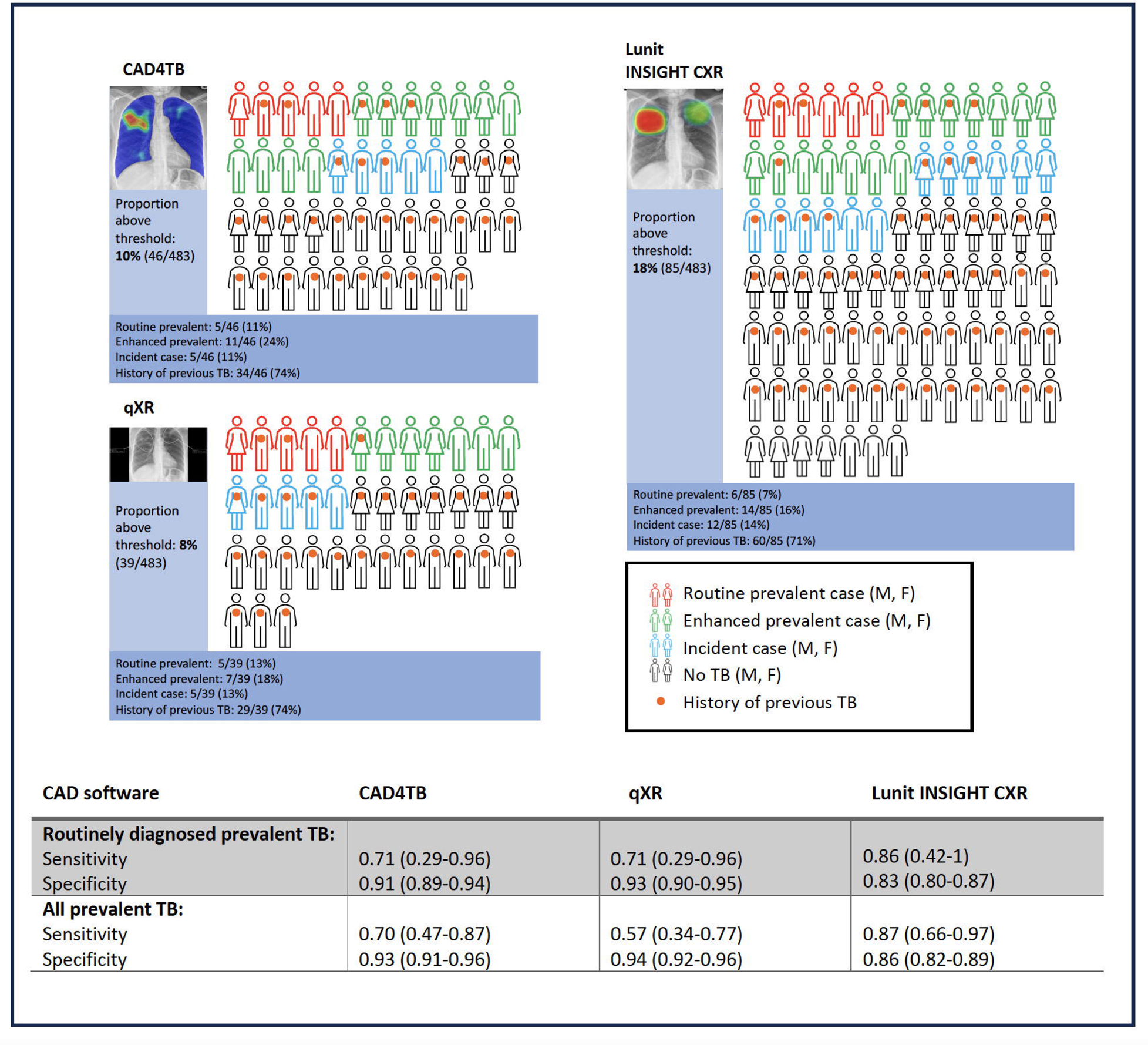
Representation of participants with baseline CAD scores for pulmonary TB above the manufacturers recommended thresholds (or in the case of CAD4TB, a commonly used threshold). Each participant with a score above recommended thresholds above which a diagnosis of TB is likely is represented as a human figure. Each figure is accurately represented as male or female, and the proportions of participants with prevalent (routine and enhanced) and incident TB are shown. Numbers represent proportion (95% confidence interval). The chest radiograph illustrations show an example of the typical output from each of the CAD products.

For each product, we calculated thresholds using the WHO TPP optimal sensitivity (0.95) and specificity (0.8) for a TB triage test for detecting any prevalent TB case. These derived thresholds were substantially lower than those recommended or currently used in practice (see Supplementary Tablbe 4)

### Comparison and combination with blood-based diagnostic tests

Of the 247 HIV uninfected participants in the *biomarker subgroup*, 245 had CRP, 247 had ESR, 242 had MTB-HR and 247 had QuantiFERON-Gold measured. Overall, 18/247 participants were diagnosed and treated for TB (6 prevalent [1 routine, 5 enhanced] and 12 with incident TB). In this subgroup, AUC of the blood-based biomarkers for all prevalent cases was lower than for CAD software – CRP 0.75 (0.55-0.96), ESR 0.78 (0.60-0.96), QFT-Gold 0.58 (0.39-0.78) and MTB-HR 0.67 (0.39-0.96) in comparison to AUC of 0.90 (0.74-1) to 0.98 (0.93-1) for the CAD software. Incorporating each blood biomarkers individually into a predictive model with CAD software did not significantly improve the AUC to detect prevalent TB (Supplementary data Table 5).

## Discussion

This is the first study reporting performance of CAD where CXR, symptom screen, routine and enhanced sputum investigation have been undertaken to screen all consenting household contacts for TB with subsequent follow-up for incident disease. We show that routine confirmatory testing with single spontaneous Xpert MTB/RIF to diagnose PTB captures only 30% (7/23) of total prevalent disease. CXR screening with CAD interpretation has excellent diagnostic performance to detect all those with prevalent disease (diagnosed routinely and with enhanced investigation) with no significant differences between the three software solutions we assessed, AUC 0.87-0.91. For all software, performance was reduced in those with previous TB and people living with HIV. Following baseline enhanced sputum investigation the performance of CAD to identify incident cases was limited (range AUC 0.60-0.65). However, using the manufacturers recommended or widely used thresholds we also showed that 30-35% of those not positive by routine (spontaneously produced) sputum Xpert MTB/RIF were either sputum positive by more intensive investigations or were diagnosed with incident TB over the follow-up period, with almost all the remainder having previous TB. Furthermore, we highlight that despite excellent AUC, the recommended CAD thresholds sensitivity is relatively low (71-86% for routine prevalent and 57-87% for all prevalent) and that thresholds could be lowered to be able to meet the WHO optimal sensitivity and/or specificity targets. Finally, we showed in a subgroup of asymptomatic HIV uninfected participants that the diagnostic performance of all CAD products to detect all prevalent TB was superior to a range of blood-based biomarkers and did not substantially improve with the addition of any of these blood based biomarkers.

Our findings suggest that the potential of CAD-CXR based screening is not being maximised as a high proportion of those above current thresholds with a negative routine confirmatory test have true TB disease that may benefit from treatment. They also show for the first time in a contemporary setting that those who have CXR changes suggestive of TB but who are bacteriologically negative by routine approaches have a similar risk of progressing to bacteriologically positive disease as shown in the recently published historical systematic review (16).

A systematic review conducted by Harris and colleagues in 2019 found three diagnostic accuracy studies that used a microbiological reference standard in screening settings (23–25). These studies examined CAD4TB only and reported sensitivities for the detection of prevalent TB ranged from 0.53 to 0.95 with specificities from 0.56 to 0.98. The authors also identified significant sources of bias, methodological limitations, and heterogeneity between the studies. More recently a prospective diagnostic accuracy study examining CAD4TB version 6, qXR version 3 and Lunit INSIGHT CXR version 3.1.0.0 was conducted by Soares and colleagues in prison settings in Brazil. They found that AUC ranged from 0.88 to 0.9 for microbiologically confirmed TB, however, excluded 70% of participants who were not able to produce spontaneous sputum samples from their primary analysis (15). Like our findings, they showed that CAD performed with greater accuracy in participants who did not have a history of previous TB. These participants are likely to have residual abnormalities visible on CXR indistinguishable from active disease.

Our study has several strengths that differentiate it from other diagnostic accuracy studies performed in screening settings: We included all household contacts who were eligible and consented for TB screening and performed thorough examination of sputum with multiple samples for investigation at baseline, we also followed participants up over a 5-year period. This enabled us to avoid misclassification of TB cases and provided a better assessment of accuracy. However, at the same time we were able to represent routine sampling to ensure wider applicability of our results. This was also the first diagnostic accuracy study to incorporate and compare a range of blood-based biomarkers, including the point-of-care Xpert MTB-HR test alongside assessment of CAD, although this analysis was restricted to a subset of HIV uninfected participants who underwent additional testing. Finally, our study was designed to minimise biases that are commonly seen in other diagnostic accuracy studies, for example, we maintained independence from CAD system operators and evaluated the software on a new dataset that had not been used in training of the software.

Our study has a number of limitations. Although we demonstrated that lower thresholds could be used and still meet WHO TPP such cut-offs were not externally validated. We screened for symptoms of TB using unexplained cough for ≥ 2 weeks, fever, night sweats and weight loss, for both HIV infected and uninfected persons rather than the recommended W4SS in HIV infection (8). This approach was taken for consistency and all participants were investigated at baseline regardless of symptoms, thus reducing the bias in our study findings. As described, this study was not powered to assess the impact of incorporating additional biomarkers on diagnostic accuracy and these need further assessment in future studies. Our study was performed in household contacts in a high burden setting where there is increased risk of re-infection over the study period, and although it is likely that many of our findings would be applicable in other screening settings, additional studies in low burden settings should be undertaken.

Following the updated WHO screening guidelines and publication of Stop TB Partnership’s Global Plan, we will continue to see scale up of CXR based screening and CAD with inevitable significant cost. Our work has shown that although CAD software is diagnostically accurate its utility is not being maximised as ∼30% of those with CAD scores above threshold with negative Xpert MTB/RIF on a spontaneous sputum sample, either have prevalent TB diagnosed by intensive sampling or develop incident TB over time. Further work is needed to establish how best to follow up and manage these patients.

## Data Availability

All data produced in the present study are available upon reasonable request to the authors

## Acknowledgments

The study was conceived by HE, SK, MR, AC and RJW. The study methods were determined by HE, LM, SK, MQ and AC. Clinical activities and data collection were undertaken by RD, RC, AT, KS, RG, NOD, AJ, ED, BS, SM, FT. Laboratory activities were undertaken by FL, MJ, NY, SG, SA and TS. The data were curated by LM, ED, AC and AJ. Analysis was perfomed by LM with supervision and support from HE, SK and MQ. The first draft of the manuscript was prepared by LM, HE and SK with all co-authors subsequently reviewing and contributing to the manuscript.

## References

1. World Health Organization. Global tuberculosis report 2019. 283 p.

2. Nguyen TBP, Nguyen TA, Luu BK, Le TTO, Nguyen VS, Nguyen KC, et al. A comparison of digital chest radiography and Xpert MTB/RIF in active case finding for tuberculosis. The International Journal of Tuberculosis and Lung Disease. 2020 Sep 1;24(9):934–40.

3. Madhani F, Maniar RA, Burfat A, Ahmed M, Farooq S, Sabir A, et al. Automated chest radiography and mass systematic screening for tuberculosis. The International Journal of Tuberculosis and Lung Disease. 2020 Jul 1;24(7):665–73.

4. Onozaki I, Law I, Sismanidis C, Zignol M, Glaziou P, Floyd K. National tuberculosis prevalence surveys in Asia, 1990–2012: an overview of results and lessons learned. Tropical Medicine & International Health. 2015 Sep 7;20(9):1128–45.

5. Law I, Floyd K, Abukaraig EAB, Addo KK, Adetifa I, Alebachew Z, et al. National tuberculosis prevalence surveys in Africa, 2008–2016: an overview of results and lessons learned. Tropical Medicine & International Health. 2020 Nov 12;25(11):1308–27.

6. Habib SS, Asad Zaidi SM, Jamal WZ, Azeemi KS, Khan S, Khowaja S, et al. Gender-based differences in community-wide screening for pulmonary tuberculosis in Karachi, Pakistan: an observational study of 311 732 individuals undergoing screening. Thorax. 2022 Mar;77(3):298–9.

7. Frascella B, Richards AS, Sossen B, Emery JC, Odone A, Law I, et al. Subclinical Tuberculosis Disease-A Review and Analysis of Prevalence Surveys to Inform Definitions, Burden, Associations, and Screening Methodology. In: Clinical Infectious Diseases. Oxford University Press; 2021. p. E830–41.

8. World Health Organisation. Module 2: Screening WHO operational handbook on tuberculosis Systematic screening for tuberculosis disease.

9. The Stop TB Partnership. THE GLOBAL PLAN TO END TB. 2023.

10. Zhen Qin Z, Ahmed S, Shahnewaz Sarker M, Paul K, Shafiq A, Adel S, et al. Can artificial intelligence (AI) be used to accurately detect tuberculosis (TB) from chest x-ray? A multiplatform evaluation of five AI products used for TB screening in a high TB-burden setting.

11. Qin ZZ, Sander MS, Rai B, Titahong CN, Sudrungrot S, Laah SN, et al. Using artificial intelligence to read chest radiographs for tuberculosis detection: A multi-site evaluation of the diagnostic accuracy of three deep learning systems. Sci Rep. 2019 Dec 1;9(1).

12. Khan FA, Majidulla A, Tavaziva G, Nazish A, Abidi SK, Benedetti A, et al. Chest x-ray analysis with deep learning-based software as a triage test for pulmonary tuberculosis: a prospective study of diagnostic accuracy for culture-confirmed disease. Lancet Digit Health. 2020 Nov 1;2(11):e573–81.

13. Harris M, Qi A, Jeagal L, Torabi N, Menzies D, Korobitsyn A, et al. A systematic review of the diagnostic accuracy of artificial intelligence-based computer programs to analyze chest x-rays for pulmonary tuberculosis. PLoS One. 2019 Sep 1;14(9).

14. Khan FA, Pande T, Tessema B, Song R, Benedetti A, Pai M, et al. Computer-aided reading of tuberculosis chest radiography: Moving the research agenda forward to inform policy. Vol. 50, European Respiratory Journal. European Respiratory Society; 2017.

15. Soares TR, Oliveira RD de, Liu YE, Santos A da S, Santos PCP dos, Monte LRS, et al. Evaluation of chest X-ray with automated interpretation algorithms for mass tuberculosis screening in prisons: A cross-sectional study. The Lancet Regional Health - Americas. 2023 Jan;17:100388.

16. Sossen B, Richards AS, Heinsohn T, Frascella B, Balzarini F, Oradini-Alacreu A, et al. The natural history of untreated pulmonary tuberculosis in adults: a systematic review and meta-analysis. Lancet Respir Med. 2023 Apr;11(4):367–79.

17. Esmail H, Coussens AK, Thienemann F, Sossen B, Mukasa SL, Warwick J, et al. High resolution imaging and five-year tuberculosis contact outcomes. medRxiv. 2023 Jul 3;

18. Bossuyt PM, Reitsma JB, Bruns DE, Gatsonis CA, Glasziou PP, Irwig L, et al. STARD 2015: an updated list of essential items for reporting diagnostic accuracy studies. BMJ. 2015 Oct 28;h5527.

19. World Health Organization. WHO consolidated guidelines on tuberculosis. Module 2: screening – systematic screening for tuberculosis disease. Geneva: World Health Organization; 2021. Licence: CC BY-NC-SA 3.0 IGO. 2021;

20. Boulle A, Heekes A, Tiffin N, Smith M, Mutemaringa T, Zinyakatira N, et al. Data Centre Profile: The Provincial Health Data Centre of the Western Cape Province, South Africa. Int J Popul Data Sci. 2019 Nov 20;4(2):1143.

21. South African Department of Health. National Tuberculosis Management Guidelines, 2014. [cited 2022 Nov 11]; Available from: https://www.tbonline.info/media/uploads/documents/national_tuberculosis_management_guidelines_%282014%29.pdf

22. World Health Organization. WHO operational handbook on tuberculosis. Module 2: screening - systematic screening for tuberculosis disease. Geneva: World Health Organization; 2021. Licence: CC BY-NC-SA 3.0 IGO. 2021.

23. Melendez J, Philipsen RHHM, Chanda-Kapata P, Sunkutu V, Kapata N, van Ginneken B. Automatic versus human reading of chest X-rays in the Zambia National Tuberculosis Prevalence Survey. The International Journal of Tuberculosis and Lung Disease. 2017 Aug 1;21(8):880–6.

24. Melendez J, Sánchez CI, Philipsen RHHM, Maduskar P, Dawson R, Theron G, et al. An automated tuberculosis screening strategy combining X-ray-based computer-aided detection and clinical information. Sci Rep. 2016;6:25265.

25. Koesoemadinata RC, Kranzer K, Livia R, Susilawati N, Annisa J, Soetedjo NNM, et al. Computer-assisted chest radiography reading for tuberculosis screening in people living with diabetes mellitus. The International Journal of Tuberculosis and Lung Disease. 2018 Sep 1;22(9):1088–94.

