## Supplementary data for "Diagnostic accuracy of Chest X-Ray Computer Aided Detection software and blood biomarkers for detection of prevalent and incident tuberculosis in household contacts followed up for 5 years"

### Members of the Imaging of TB household contacts group

| First name and Inital | Surname | Affiliation |
| --- | --- | --- |
| Clifton E. | Barry III | Tuberculosis Research Section, Laboratory of Clinical Immunology and Microbiology, National Institute of Allergy and Infectious Diseases, National Institutes of Health, Bethesda, MD. |
| Jerrold J. | Ellner | Rutgers- New Jersey Medical School, Center for Emerging Pathogens, Newark, NJ, United States. |
| JoAnne L. | Flynn | Department of Microbiology and Molecular Genetics, University of Pittsburgh School of Medicine, PA 15261, United States of America |
| Torben | Heinsohn | Department of Epidemiology, Helmholtz Centre for Infection Research, Braunschweig, Germany. |
| C. Robert | Horsburgh Jr. | Department of Medicine, Boston University School of Medicine, Boston, MA, USA; Departments of Epidemiology, Biostatistics and Global Health, Boston University School of Public Health, Boston, MA, USA |
| Karen R. | Jacobson | Department of Medicine, Boston University School of Medicine, Boston, MA, USA; Departments of Epidemiology, Biostatistics and Global Health, Boston University School of Public Health, Boston, MA, USA |
| Stephanus T. | Malherbe | Department of Science and Technology/National Research Foundation Centre of Excellence in Biomedical Tuberculosis Research, South African Medical Research Council for Tuberculosis Research, Division of Molecular Biology and Human Genetics, Department of Biomedical Sciences, Faculty of Health Sciences, Stellenbosch University, Cape Town, South Africa. |
| Padmini | Salgame | Rutgers- New Jersey Medical School, Center for Emerging Pathogens, Newark, NJ, United States. |
| Dylan | Sheerin | The Walter and Eliza Hall Institute of Medical Research, Parkville, Victoria 3052, Australia Department of Medical Biology, University of Melbourne, Parkville, 3052, Australia. |
| Elizabeth | Streicher | Department of Science and Technology/National Research Foundation Centre of Excellence in Biomedical Tuberculosis Research, South African Medical Research Council for Tuberculosis Research, Division of Molecular Biology and Human Genetics, Department of Biomedical Sciences, Faculty of Health Sciences, Stellenbosch University, Cape Town, South Africa. |
| Mpho | Tlala | Department of Science and Technology/National Research Foundation Centre of Excellence in Biomedical Tuberculosis Research, South African Medical Research Council for Tuberculosis Research, Division of Molecular Biology and Human Genetics, Department of Biomedical Sciences, Faculty of Health Sciences, Stellenbosch University, Cape Town, South Africa. |
| Laura E. | Via | Tuberculosis Research Section, Laboratory of Clinical Immunology and Microbiology, National Institute of Allergy and Infectious Diseases, National Institutes of Health, Bethesda, MD Tuberculosis Imaging Program, Division of Intramural Research, National Institute of Allergy and Infectious Diseases, National Institutes of Health, Bethesda, MD. |
| Gerhard | Walzl | Department of Science and Technology/National Research Foundation Centre of Excellence in Biomedical Tuberculosis Research, South African Medical Research Council for Tuberculosis Research, Division of Molecular Biology and Human Genetics, Department of Biomedical Sciences, Faculty of Health Sciences, Stellenbosch University, Cape Town, South Africa. |
| Robin | Warren | Department of Science and Technology/National Research Foundation Centre of Excellence in Biomedical Tuberculosis Research, South African Medical Research Council for Tuberculosis Research, Division of Molecular Biology and Human Genetics, Department of Biomedical Sciences, Faculty of Health Sciences, Stellenbosch University, Cape Town, South Africa. |
| James | Warwick | Division of Nuclear Medicine, Department of Medical Imaging and Clinical Oncology, Stellenbosch University, Cape Town, South Africa |

### Details of household contact recruitment

Index cases aged ≥15 years with at least rifampicin resistant pulmonary tuberculosis (TB) confirmed by Xpert MTB/RIF or culture consented to a household visit and to contact their household members and close contacts. Household contacts (HHC) were defined as individuals sleeping in the same dwelling or room and/or providing themselves jointly with food or other essentials for living during the day with the index case for at least 7 days during the 3 months prior to the index case being diagnosed with TB. All HHC ≥18 years were offered an appointment at the study clinic for TB screening. Written, informed consent was obtained from all index cases and HHC participants.

### Inclusion and exclusion criteria – n=250 biomarker subgroup:

*Inclusion Criteria*

1. Household contact of DR-TB index case
2. Age 18 years or over
3. Consents to participating in study
4. Willing to undergo HIV counselling and testing (HCT)

*Exclusion Criteria*

1. HIV infection
2. On TB treatment at the time of screening
3. Symptoms or signs of active TB
4. Symptoms or signs of acute illness
5. Age >65 years
6. Smoker >30 pack years
7. Diagnosis of malignancy
8. Diagnosis of chronic lung infection other than TB (e.g., non-tuberculosis mycobacteria [NTM], Fungal)
9. Diagnosis of chronic inflammatory condition associated with pulmonary pathology (e.g., Sarcoidosis, Rheumatoid Arthritis, Wegener’s granulomatosis, bronchiectasis)
10. Inhaled or systemic steroid use within previous 2 weeks
11. Breast feeding, pregnant, or planning pregnancy over next 3 months
12. Unable to be followed up for 6 months
13. Uncontrolled diabetes mellitus

### Xpert MTB-HR Methodology

***Xpert MTB-HR Tempus Tube validation cohort***

The Xpert Mtb-HR cartridge is designed to be used with 100 μl of capillary finger prick blood or pelleted Paxgene blood cells resuspended in TB HR Lysis Buffer added directly to the cartridge. To validate its use with stored Tempus tube blood a comparison was performed between results obtained using fresh capillary blood and stored Tempus tubes taken from 49 individuals. Participants included 19 individuals newly diagnosed with TB at the Site B Ubuntu clinic in Khayelitsha (6 HIV-infected, 13 HIV uninfected) and 30 healthy controls, confirmed not to have TB recruited via the Site B HIV wellness clinic, including 12 HIV infected individuals previously established on ART and 18 individuals confirmed HIV uninfected following HIV counselling and testing, performed by the wellness clinic staff. Written informed consent was obtained from all participants following human research ethical approval received from University of Cape Town Faculty of Health Sciences (449/2014) and University College London (19219/001).

***Xpert MTB-HR Tempus Tube vs capillary blood comparison***

Finger prick blood was taken using a Minivette (BD) collecting 100 μl of whole blood that was added directly to the Xpert MTB-HR cartridge within 15 min of blood draw. Following the fingerpick, venus blood was also collected in a Tempus Tube that was stored at -80^o^C (for > 7 days until processed) and an EDTA tube that was stored at RT on the day until the capillary MTB-HR result was confirmed valid. If an invalid result was recorded the test was repeated adding 100 μl of EDTA whole blood directly to a new cartridge. To run the Tempus tubes, following overnight thawing at 4^o^C, tubes were inverted 2-3 times and 380 μl of Tempus blood transferred to 1.5 ml Eppendorf and centrifuged at 3000g for 5 min. Supernatant was removed, cell pellet resuspended in 100 μl of TB HR Lysis Buffer (Cepheid). Samples were vortexed for 10 sec to dissolve the pellet and then 100 μl transferred to the Xpert MTB-HR cartridge. All cartridges were run within 30 min of blood addition on a GeneXpert instrument using the TB Host Response Alpha software module. Results indicated no significant difference in fresh blood vs stored Tempus blood results for the HIV-infected and HIV-uninfected controls and only a small decrease in median LDA (fresh 0.48 vs Tempus 0.30, p=0.029) for the TB group (A below). Based on the LDA threshold cutoff of <2 indicating TB, 16/19 (84%) TB patients had LDA <2 for fresh blood and 17/19 (89%) for Tempus blood. All stored Tempus tubes from HHC in the biomarker substudy were therefore run as detailed above.

***Xpert MTB-HR Tempus Tube vs extracted RNA comparison***

Of the 247 HHC in the biomarker substudy that had 2 Tempus tubes stored, 44 had already had RNA extracted from both Tempus tubes and had no Tempus tubes remaining. To validate equivalent results using extracted RNA from the same Tempus tube, one Tempus tube from each of 9 individuals with 2 remaining tubes were thawed and first the Tempus tube blood analysed on the Xpert MTB-HR cartridge, then RNA extracted from the same Tempus tube using the Norgen Preserved Blood RNA kits (for Tempus Tubes), following manufacturers protocol including DNAse treatment (Norgen). Three volumes of RNA to be added to the cartridge were then tested resuspending to a total of 100 μl using TB HR Lysis Buffer: 2 μl, 1,8 μl and 0.5 μl. Results indicated a small significant higher LDA value using 2 μl and no difference using 0.5 μl (B below). Given the increased potential of pipetting error when adding 0.5 μl, 1 μl + 99 μl TB HR Lysis Buffer was used to run the 44 samples with only RNA remaining.

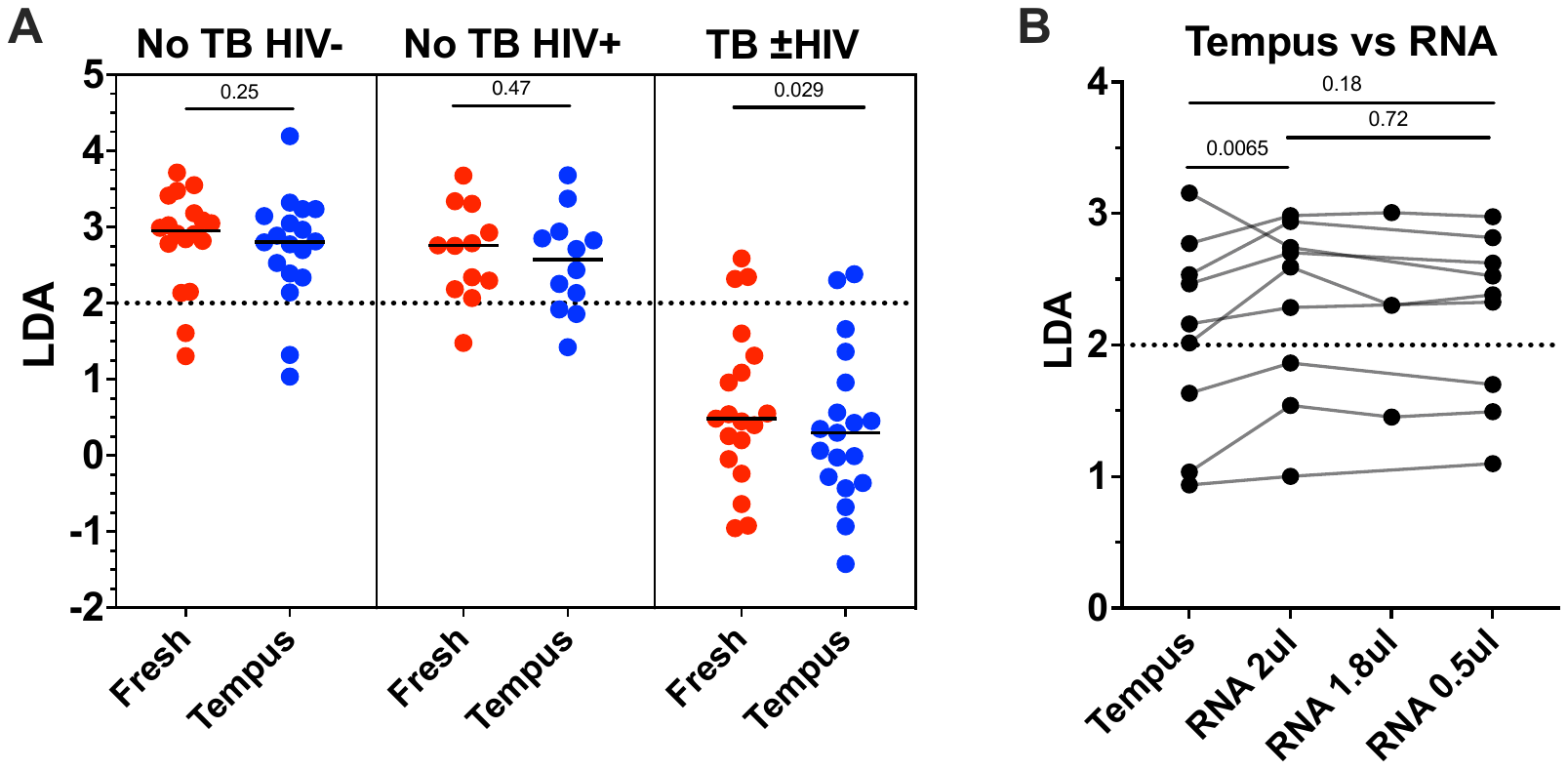

**Validation of using Tempus tube blood and extracted RNA in the Xpert MTB-HR cartridge**. (**A**) LDA results from asymptomatic HIV-uninfected (n=18), asymptomatic HIV-infected (n=12) no TB controls and symptomatic newly diagnosed TB patients (n=19), comparing results between fresh blood and stored Tempus tube blood. Median indicated by line, analysed by Wilcoxon matched-pairs test. (**B**) LDA results from asymptomatic HIV-uninfected (n=9) person comparing using Tempus tube blood and different volumes of RNA extracted from the same Tempus tube. Friedman test with Dunn's multiple comparison testing.

### Additional CAD methodology

CAD software was installed by the companies on local servers (one for each software) that are under the control of FIND, one of the partners in this study. After successful installation, access to the servers was lifted for the CAD vendors to allow for an independent evaluation. Anonymised digital images were uploaded via a secure server to FIND and then processed by the CAD software. No images were shared with the software manufacturers. The CAD output consists of a score along a continuous scale (CAD4TB and Lunit INSIGHT CXR 0-100, qXR 0-1), where high scores indicate a higher probability of active TB.

Both qXR and Lunit INSIGHT CXR come with manufacturer recommended threshold scores for TB (0.5 and 15 respectively), above which the X-ray is considered likely compatible with TB and below which it is not. CAD4TBv7 has a default threshold of 60, but in practice many different thresholds are used as the developers encourage and support users to find an adjusted thresholds most appropriate for their setting. In practice, this cut off ranges between ~50-70 and we used the lower score of 50 for this analysis.

### Supplementary Table 1 : Microbiology results for participants with prevalent TB (n=23)

| **Smear status:**  Scanty  1+  2+  3+ | 2 (9%)  2 (9%)  5 (22%)  1 (4%) |
| --- | --- |
| *Mtb* detected by Xpert MTB/RIF in at least 1 baseline sample | 14 (61%) |
| Proportion with any culture positive sample | 20 (87%) |

### Supplementary Figure 1: AUC ROCs for incident TB over time for each CAD software.

CAD4TB

qXR

Lunit INSIGHT CXR

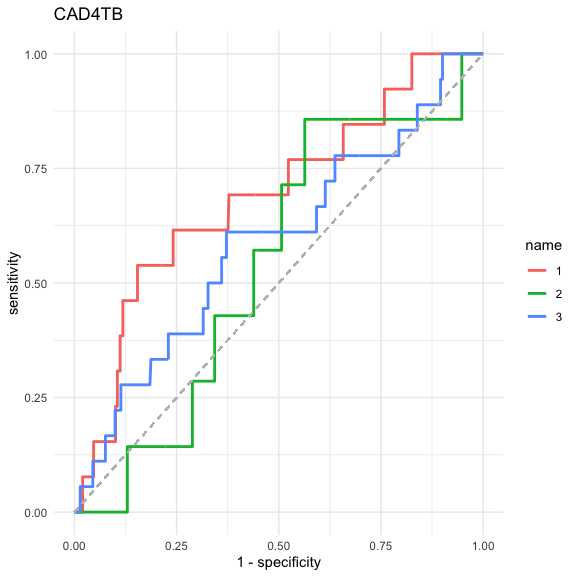

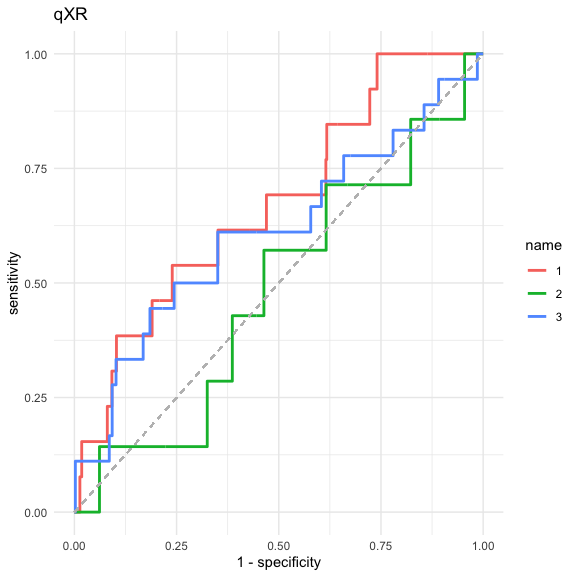

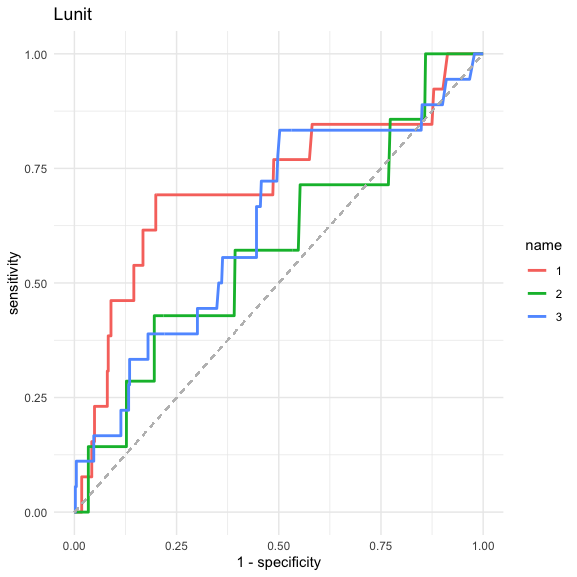

>0 - ≤12 months

>12 - ≤24 months

>24 months

|  | AUC ROC for CAD4TB (95% CI) | AUC ROC for qXR (95% CI) | AUC ROC for Lunit INSIGHT CXR (95% CI) |
| --- | --- | --- | --- |
| Incident TB from 1-12 months **(n= 460)** | 0.69 (0.53-0.85) | 0.67 (0.52-0.82) | 0.71 (0.54-0.89) |
| Incident TB from 13-24 months **(n= 447)** | 0.543 (0.35-0.74) | 0.48 (0.25-0.71) | 0.58 (0.34-0.82) |
| Incident TB from 25 months **(n= 440)** | 0.61 (0.45-0.77) | 0.58 (0.44-0.73) | 0.63 (0.49-0.77) |

### Supplementary Table 2: Subgroup analysis - AUC ROC for participants with and without HIV infection and for those with and without a previous history of TB.

|  | **Previous TB (n=109)** | **No previous TB (n=374)** | ***P value*** |
| --- | --- | --- | --- |
| **AUC for all prevalent TB**  CAD4TB  qXR  Lunit INSIGHT CXR | 0.65 (0.43-0.87)  0.69 (0.54-0.85)  0.70 (0.57-0.83) | 0.94 (0.84-1)  0.90 (0.77-1)  0.93 (0.82-1) | *0.02*  *0.04*  *0.01* |
| **AUC for all incident TB**  CAD4TB  qXR  Lunit INSIGHT CXR | 0.50 (0.31-0.69)  0.49 (0.28-0.71)  0.54 (0.33-0.74) | 0.62 (0.50-0.74)  0.65 (0.52-0.77)  0.69 (0.56-0.81) | *0.31*  *0.21*  *0.23* |

|  | **HIV infected (n=136)** | **HIV uninfected (n=340)** | ***P value*** |
| --- | --- | --- | --- |
| **AUC for all prevalent TB**  CAD4TB  qXR  Lunit INSIGHT CXR | 0.78 (0.59-0.96)  0.85 (0.68-1)  0.85 (0.69-1) | 0.94 (0.90-0.98)  0.91 (0.82-0.997)  0.96 (0.93-0.99) | *0.09*  *0.5*  *0.18* |
| **AUC for all incident TB**  CAD4TB  qXR  Lunit INSIGHT CXR | 0.63 (0.51-0.76)  0.63 (0.50-0.76)  0.64 (0.49-0.79) | 0.57 (0.41-0.73)  0.64 (0.48-0.79)  0.66 (0.52-0.80) | *0.55*  *0.95*  *0.84* |

### Supplementary Table 3: The sensitivity and specificity of each CAD software using the manufacturer recommended (or commonly used) threshold.

|  | CAD4TB | | | qXR | | | Lunit INSIGHT CXR | | |
| --- | --- | --- | --- | --- | --- | --- | --- | --- | --- |
|  | Routine prevalent TB cases | All prevalent TB cases | All TB cases | Routine prevalent TB cases | All prevalent TB cases | All TB cases | Routine prevalent TB cases | All prevalent TB cases | All TB cases |
| Threshold | **50** | | | **0.5** | | | **15** | | |
| Sensitivity | 0.71 (0.29-0.96) | 0.70 (0.47-0.87) | 0.34 (0.23-0.48) | 0.71 (0.29-0.96) | 0.57 (0.34-0.77) | 0.52 (0.39-0.65) | 0.86 (0.42-0.996) | 0.87 (0.66-0.97) | 0.52 (0.39-0.65) |
| Specificity | 0.91 (0.89-0.94) | 0.93 (0.91-0.96) | 0.94 (0.91-0.96) | 0.93 (0.90-0.95) | 0.94 (0.92-0.96) | 0.98 (0.97-0.99) | 0.83 (0.80-0.87) | 0.86 (0.82-0.89) | 0.87 (0.84-0.90) |
| PPV | 0.11 (0.07-0.17) | 0.35 (0.26-0.45) | 0.46 (0.33-0.58) | 0.13 (0.08-0.21) | 0.33 (0.23-0.46) | 0.82 (0.68-0.91) | 0.07 (0.05-0.10) | 0.24 (0.19-0.29) | 0.38 (0.30-0.46) |
| NPV | 0.995 (0.99-0.999) | 0.98 (0.97-0.99) | 0.91 (0.89-0.92) | 0.995 (0.986-0.999) | 0.98 (0.96-0.99) | 0.93 (0.92-0.95) | 0.998 (0.98-0.999) | 0.99 (0.98-0.997) | 0.93 (0.90-0.94) |
| TP | 5 | 16 | 21 | 5 | 13 | 32 | 6 | 20 | 32 |
| FP | 41 | 30 | 25 | 35 | 27 | 70 | 79 | 65 | 53 |
| TN | 435 | 430 | 397 | 441 | 433 | 352 | 397 | 395 | 369 |
| FN | 2 | 7 | 40 | 2 | 10 | 29 | 1 | 3 | 29 |

PPV = positive predictive value, NPV = negative predictive value

TP = True positive, FP = False positive, TN = True negative, FN = False negative.

### Supplementary Table 4: Thresholds derived from the WHO target product profile optimal sensitivity (0.95) and specificity (0.8) for a TB triage test.

Using a sensitivity of 0.95 the derived thresholds were 3.36 (specificity 0.22), 0.0097 (specificity 0.46), and 5.11 (specificity 0.81) for CAD4TB, qXR and Lunit INSIGHT CXR respectively. Using a specificity of 0.8 the derived thresholds were 25.6 (sensitivity 0.83), 0.038 (sensitivity 0.87), 4.47 (sensitivity 0.96) for CAD4TB, qXR and Lunit INSIGHT CXR respectively. These data suggest that current thresholds used by manufacturers could potentially be lowered in a screening setting where more intensive sputum sampling approaches are taken.

|  | CAD4TB | | qXR | | Lunit INSIGHT CXR | |
| --- | --- | --- | --- | --- | --- | --- |
|  | All prevalent TB cases | | All prevalent TB cases | | All prevalent TB cases | |
| **Threshold** | **3.36** | **25.6** | **0.0097** | **0.038** | **5.11** | **4.47** |
| Sensitivity | 0.95 (fixed) | 0.83 | 0.95 (fixed) | 0.87 | 0.95 (fixed) | 0.96 |
| Specificity | 0.22 | 0.8 (fixed) | 0.46 | 0.8 (fixed) | 0.81 | 0.8 (fixed) |
| PPV | 0.06 | 0.17 | 0.08 | 0.18 | 0.20 | 0.19 |
| NPV | 0.99 | 0.99 | 0.995 | 0.99 | 0.997 | 0.997 |
| TP | 22 | 19 | 22 | 20 | 22 | 22 |
| FP  Incident cases | 361  32 (8.9%) | 91  14 (15.3%) | 250  24 (9.6%) | 92  15 (16.3%) | 87  16 (18.4%) | 92  16 (17.4%) |
| TN | 99 | 369 | 210 | 368 | 373 | 368 |
| FN | 1 | 4 | 1 | 3 | 1 | 1 |

PPV = positive predictive value, NPV = negative predictive value

TP = True positive, FP = False positive, TN = True negative, FN = False negative.

Incident cases = number (percentage) of cases considered false positive that subsequently develop incident TB over follow-up

### Supplementary Table 5: AUC ROC for each CAD software for detecting prevalent and incident TB, in combination with biomarkers:

|  | **All prevalent TB cases** | **Incident TB cases** |
| --- | --- | --- |
| CRP (n=245) | 0.75 (0.55-0.96) | 0.59 (0.45-0.73) |
| ESR (n=249) | 0.78 (0.60-0.96) | 0.61 (0.46-0.76) |
| QuantiFERON (TBA) (n=247) | 0.58 (0.39-0.78) | 0.55 (0.38-0.72) |
| Host response (n=242) | 0.67 (0.39-0.96) | 0.59 (0.40-0.79) |
| CAD4TB  CAD4TB + CRP  CAD4TB + ESR  CAD4TB + QuantiFERON  CAD4TB + HR | 0.95 (0.89-1)  0.94 (0.87-1)  0.94 (0.83-1)  0.98 (0.95-0.999)  0.93 (0.83-1) | 0.64 (0.42-0.85)  0.66 (0.43-0.89)  0.73 (0.58-0.89)  0.70 (0.50-0.90)  0.77 (0.62-0.92) |
| qXR  qXR + CRP  qXr + ESR  qXR + QuantiFERON  qXR + HR | 0.90 (0.74-1)  0.90 (0.73-1)  0.88 (0.65-1)  0.98 (0.95-0.998)  0.86 (0.59-1) | 0.68 (0.47-0.89)  0.71 (0.50-0.93)  0.77 (0.65-0.90)  0.71 (0.50-0.92)  0.82 (0.70-0.93) |
| Lunit INSIGHT CXR  Lunit INSIGHT CXR + CRP  Lunit INSIGHT CXR + ESR  Lunit INSIGHT CXR + QuantiFERON  Lunit INSIGHT CXR + HR | 0.98 (0.93-1)  0.93 (0.80-1)  0.90 (0.70-1)  0.995 (0.99-1)  0.89 (0.67-1) | 0.72 (0.54-0.91)  0.78 (0.61-0.95)  0.78 (0.65-0.90)  0.71 (0.52-0.91)  0.82 (0.71-0.93) |

### Supplementary Figure 2: CAD score and host response blood test result for 247 HIV uninfected, asymptomatic participants for each CAD software.

***Host-response score by CAD score for each CAD software:*** *Each dot represents a study participant. The horizontal blue line represents the threshold* ***below*** *which the host-response test is positive, the vertical blue line represents the manufacturer recommended threshold (or in the case of CAD4TB commonly used in the field) above which is consistent with radiographic TB.*

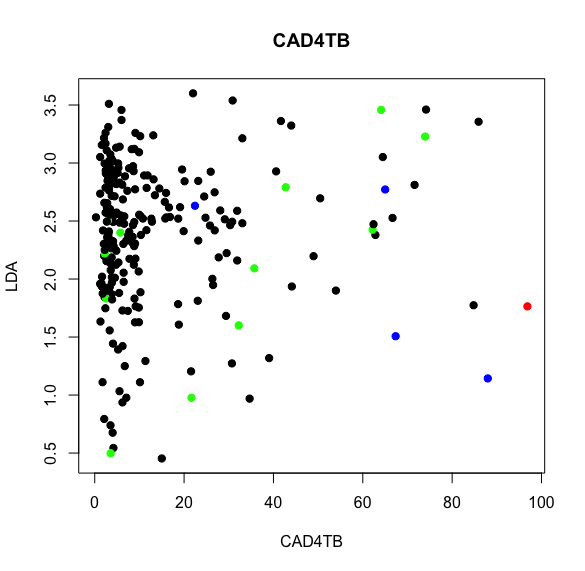

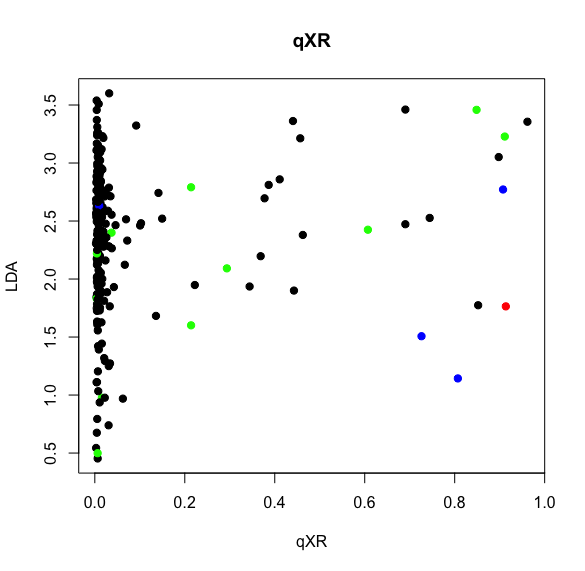

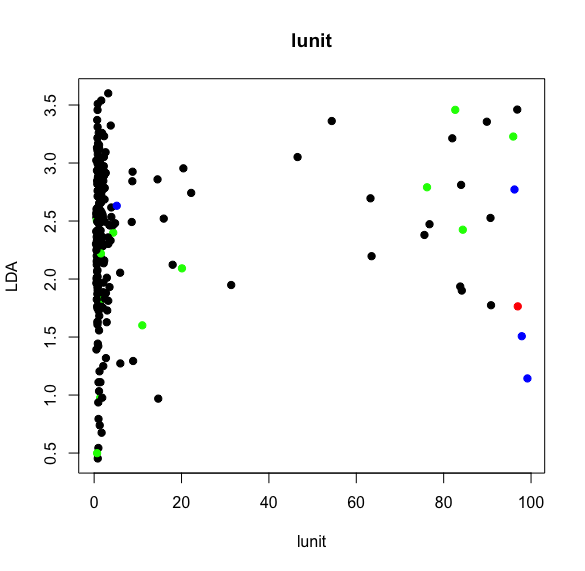

***= Routine prevalent TB case***

***= Enhanced prevalent TB case***

***= Incident TB case***

***= no TB***

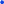

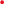

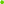

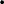

Lunit INSIGHT CXR
